# Genetically determined platelet traits impact stroke risk through multiple mechanisms and cell types

**DOI:** 10.1101/2025.08.01.25332822

**Authors:** Kim Ha, Katherine Hartmann, Renae L Judy, Michael Levin, Scott Damrauer, Christopher S Thom

## Abstract

Stroke remains a leading cause of death and disability worldwide. Current antiplatelet and anticoagulant treatments are prone to failure. Heritable blood and platelet traits contribute to stroke risk, but related mechanisms are not fully understood. Platelets bind to damaged endothelial walls to initiate thrombosis, but erythrocyte and leukocyte recruitment are involved in stroke pathogenesis. We aimed to identify causal blood-related cells and mechanisms that modulate stroke risk. By two sample Mendelian Randomization (MR), increased platelet count heightened stroke risk (Odds ratio [OR] 1.03 per 1 SD unit increase in platelet count, P=1×10^-2^). However, these effects were relatively weak and complicated by similar effects from erythrocyte and leukocyte traits. To ascertain key blood traits that influence stroke, we applied Bayesian Model Averaging (MR-BMA) and identified platelet count and mean platelet volume as key positive regulators for stroke risk. We validated an epidemiologic association between increased platelet count and higher stroke risk among a large patient cohort. Taken together, these findings indicate that platelet traits are the most critical risk factors for stroke, among analyzed blood cell traits. To deconvolute multiple underlying genetic mechanisms by which platelet traits impact stroke risk, we clustered platelet count variants using noise-augmented directional clustering (NAvMix). We identified 13 clusters, two of which were highly predictive for increased stroke risk (OR 1.31 per SD unit increase in platelet count, P<1×10^-6^). Pathway analyses on eQTLs linked to variants in these subclusters indicated enrichment for endothelial cell adhesion or platelet reactivity. Colocalization analysis of stroke and platelet count loci identified genes implicated in platelet reactivity (*RIN3*) and peroxisome biogenesis (*PEX6/PEX29*). These findings reflect complex mechanisms underlying platelet trait variation and reveal key pathways that influence stroke risk through multiple cell types and biological mechanisms, including platelet biology and endothelial cell adhesion. An approach combining novel MR methods with subclustering may be a viable method to ascertain causal mechanisms related to other closely related exposure traits.

## Introduction

Stroke is a major cause of mortality and long-term disability worldwide^1^. Strokes occur when a cerebral blood vessel is blocked (ischemic) or ruptured (hemorrhagic), resulting in motor, visual, sensory and/or cognitive deficits. Strokes can be subtyped into large artery stroke (LAS), small vessel stroke (SVS), and cardioembolic stroke (CES) depending on the origin and location of the initial clot^2^. Stroke prevention and treatment has focused on modifiable risk factors, such as obesity, hypertension, hypercholesterolemia, tobacco use, and physical inactivity^3^. Antiplatelet and anticoagulant therapy are also standard of care for stroke prevention and treatment. The high population burden of stroke despite existing measures demonstrates variable effectiveness in preventive measures, let alone the inability to avoid disease progression, recurrence, and mortality^4^. Heterogeneity in stroke etiology underscores the need to define additional causal mechanisms in stroke pathophysiology to identify targeted therapies for stroke prevention and treatment.

Blood cells play central roles in thrombosis, inflammation, and clot structure that contribute to stroke risk and severity. Multiple blood cell lineages contribute to stroke pathology. In many cases, it is difficult to parse specific contributions of individual blood cell types. Platelets are essential mediators of thrombosis, initiating coagulation cascades upon vascular injury. Stroke prevention strategies target platelet activation, aggregation, and coagulation to reduce stroke risk. However, white blood cells can initiate inflammation that activate platelets and, in turn, platelets recruit leukocytes to amplify inflammation^5–7^. Red blood cells influence clot formation by modulating platelet reactivity and blood viscosity, with emerging roles in stabilizing fibrin networks that facilitate clot maturation and contraction^8^.

Observational and genetic studies have linked increased platelet, white blood cell, and red blood cell counts with heightened cardiovascular disease risks^9–13^. However, clinical observational studies can be susceptible to confounding^14^ and prior genetic studies lacked power to resolve some trait relationships, as reflected in surprisingly weak effect sizes^15^. We hypothesized that shared genetic architecture across blood cell traits complicated these efforts to pinpoint traits that directly influence stroke risk. For example, all blood cells differentiate from common hematopoietic stem and progenitor cells (HSPCs). Genetic influences on HSPCs can be reflected in similar effects on all cellular progeny^16,17^. Alternatively, impacts on mature blood cells can result in genetic influences on a single blood cell lineage (e.g., only platelets or erythrocytes)^18^.

Mendelian Randomization (MR) is a causal inference framework that uses genetic variants as instruments to assess whether risk factors (e.g., higher blood counts) causally influence outcomes (e.g., stroke) provided that certain conditions are met^19^. Extensions of the classical two sample MR and multivariable MR (MVMR) methods, including MR-BMA (Bayesian Model Averaging), can more readily discriminate individual causal traits from a group of genetically correlated exposures^20^. Separately, clustering methods have been devised to parse multiple mechanisms that underlie a given trait. For example, Noise-Augmented Directional Clustering (NAvMix) clusters variants based on proportional associations with related traits^21^. Together, MR-BMA and NAvMix offer powerful approaches to resolve the effects of related exposures into distinct biological pathways and mechanisms.

We designed this study to leverage novel genetic epidemiology approaches to elucidate potential mechanisms linking blood cell traits and stroke risk. We sought to 1) evaluate the potential causal contribution of platelet and other blood traits to stroke risk using genetic causal inference approaches, 2) confirm epidemiologic associations between relevant blood traits and stroke risk, and 3) resolve specific pathways and mechanisms linking blood trait variation to stroke susceptibility.

## Results

### Genetically influenced blood count variation impacts stroke risk

We conducted two-sample Mendelian Randomization (MR) to test if genetically influenced variation in 15 blood traits impacts stroke risk, using the largest GWAS to date (**Fig. 1**). We identified between 280 and 1,588 linkage-independent single nucleotide polymorphisms (SNPs) per blood trait, which formed well-powered instrumental variables for MR studies. By the inverse-variance weighted (IVW) method, we observed significant positive associations for platelet count, erythroid, and leukocyte counts on stroke risk and most stroke subtypes (**Fig. 1**). Increased platelet count was associated with all strokes (OR 1.030 [95% CI: 1.007 to 1.054], P = 9.9×10^-3^), ischemic stroke, and cardioembolic stroke (**Fig. 1**). Increased hemoglobin level (HGB) and other erythroid traits were also linked with an increased risk of all strokes (OR 1.050 [1.015 to 1.087], P = 4.6×10^-3^) and ischemic stroke subtypes (**Fig. 1**). Leukocyte traits, including lymphocyte count (LYM) and white blood cell count (WBC), were also positively associated with all stroke and ischemic stroke risks (OR 1.039 [1.007 to 1.071] per SD unit increased in WBC, P = 1.5×10^-2^; OR 1.044 [1.014 to 1.075 SD units] per SD unit increase in LYM, P = 3.7×10^-3^).

**Figure 1.**
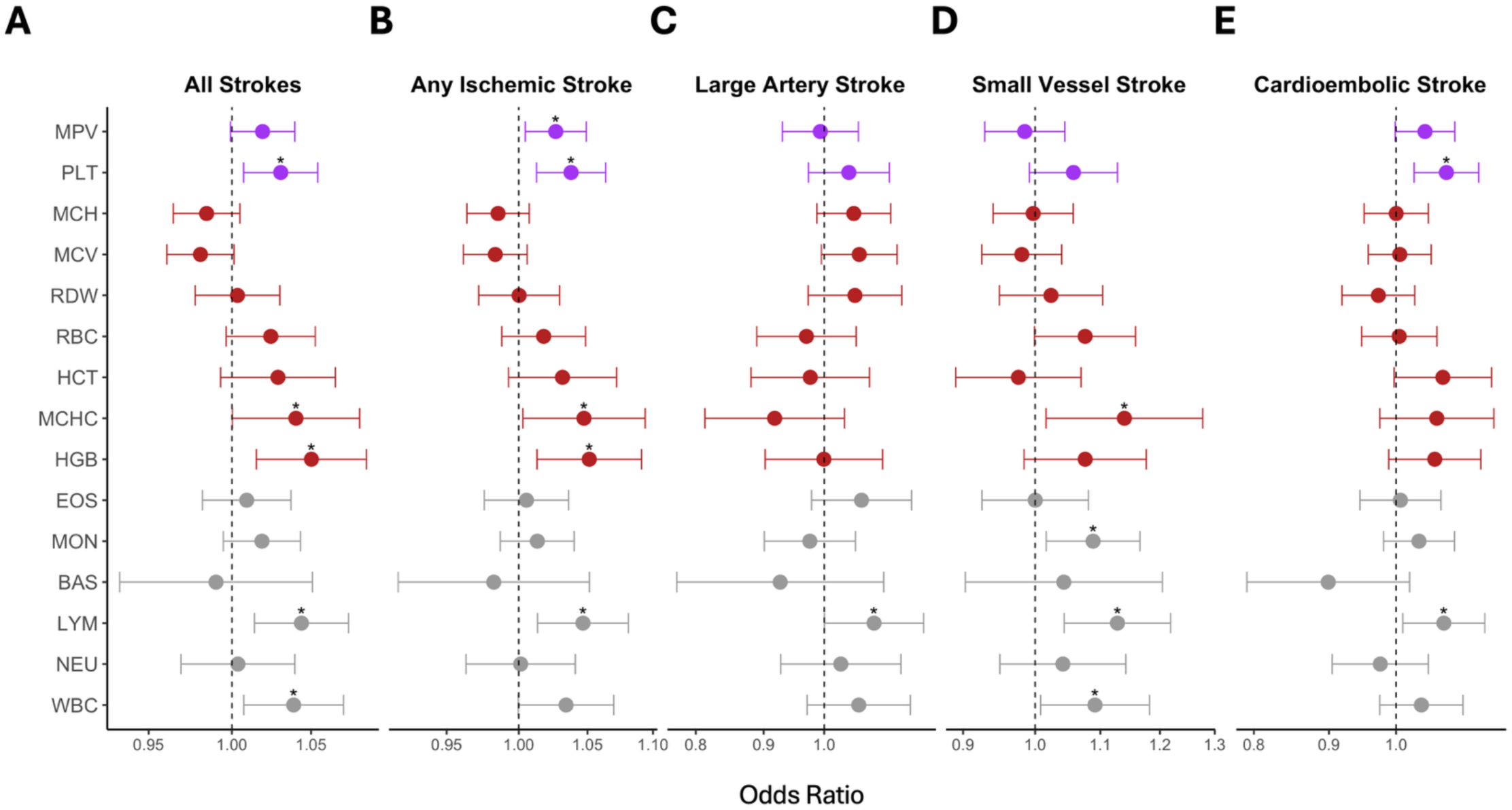
Mendelian Randomization (MR) effect estimates for quantitative blood traits on stroke risk. Forest plots of MR estimates (inverse variance weighted (IVW) method) for 15 quantitative blood traits on the risk of (A) all strokes, (B) any ischemic stroke, (C) large artery stroke, (D) small vessel stroke, or (E) cardioembolic stroke. Purple indicates platelet traits, red indicates erythroid traits, and gray indicates leukocyte traits. Odds ratios are shown per 1 standard deviation (SD) units pf blood traits with bars indicating 95% confidence intervals. *P < 0.05.

These findings suggest contributions from multiple blood cell lineages to stroke risk, with variable effects across stroke subtypes indicating distinct mechanisms. On aggregate, the effect sizes appeared modest compared to what should be robust associations given that blood cells directly instigate and propagate thrombosis and stroke pathophysiology. However, these MR experiments could not discriminate effects of correlated blood traits or prioritize key blood cell types, which limits interpretation of these causal genetic relationships.

### Platelet trait variation is most closely linked to stroke risk compared to other blood lineages

We hypothesized that parsing the effects of blood traits would reveal key cell types that differentially impact stroke risk. MR Bayesian model averaging (MR-BMA) is a novel multivariable MR approach designed to resolve independent contributions of correlated exposure traits to identify the most influential factors^20^. This approach accounts for genetic correlation and estimates a marginal inclusion probability (MIP) for each exposure trait. We applied MR-BMA to an IV comprising 4,266 SNPs that were significantly associated with variation in one or more blood traits^23^. Platelet count (PLT) and mean platelet volume (MPV) were the top-ranked traits associated with increased stroke risk (PLT MIP = 0.975, MPV MIP = 0.930, **Fig. 2**). We also observed a high MIP for hemoglobin (HGB MIP = 0.912). Increased PLT, MPV, and HGB each heightened stroke risk in this experiment (PLT effect = 0.075, MPV effect = 0.048, HGB effect = 0.107). Our MR-BMA findings highlighted the critical importance of genetically influenced platelet biology in mitigating stroke risk. We therefore chose to focus subsequent analyses on platelet loci to elucidate biological pathways by which platelet biology impacts stroke risk.

**Figure 2.**
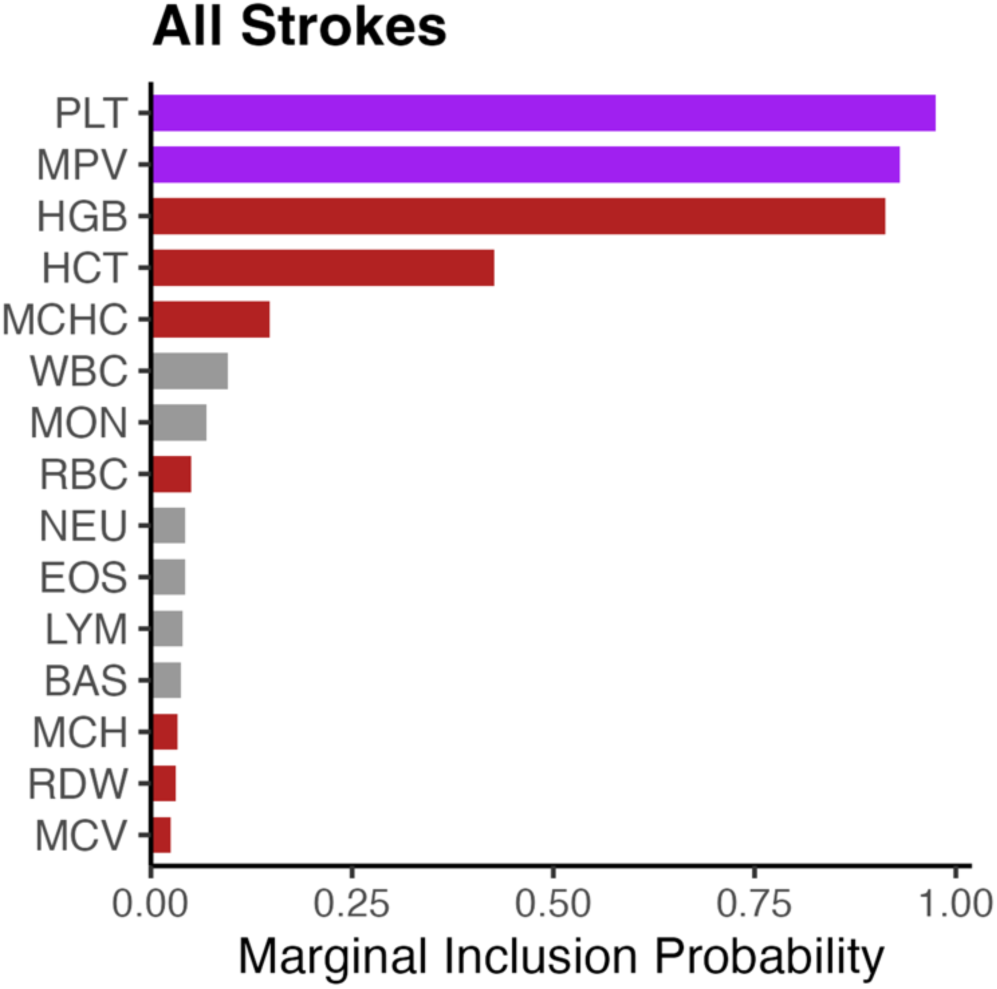
MR-BMA marginal inclusion probability estimates for 15 quantitative blood traits in stroke risk models. Bars represent the marginal inclusion probability (MIP) of each blood trait in models predicting stroke risk. MIP is defined as the sum of posterior probabilities over all models in which the trait is included. Platelet quantitative traits (PLT and MPV) were prioritized with the highest MIPs, followed by hemoglobin (HGB). Colors indicate trait categories (purple for platelet traits, red for erythroid traits, gray for leukocyte traits).

### Increased platelet count variation is epidemiologically associated with stroke risk

Antiplatelet strategies are effective in limiting stroke risk^25^, but there is limited epidemiologic evidence linking platelet count with this effect^9^. To confirm relationships between platelet count and the risks of stroke or mortality, we analyzed data from the electronic health records of 324,677 subjects (**Tables 1-2)**. We excluded 4,759 (1.4%) with thrombocytopenia and 4,973 (1.5%) with thrombocytosis. The mean follow-up time for subjects was 9.04 years, during which 38,174 (12%) participants died. Platelet count was stable among subjects who had multiple platelet counts obtained within the study period (absolute change 2.19 x 10^6^ platelets/ml, n = 38). The median platelet count among included subjects was 2.26 (1^st^-3^rd^ quartile 1.89-2.69, **Table 2**). Platelet count was higher among women, 2.36 (1^st^-3^rd^ quartile 1.98-2.79) than men, 2.10 (1^st^-3^rd^ quartile 1.76-2.50, p < 2.2×10^-16^). Age was negatively correlated with platelet count (Spearman’s rho -0.11, p < 2.2×10^-16^).

**Table 1.** Clinical characteristics for the UPHS population.

| **Characteristic** | **N = 324,778<sup>1</sup>** |
| --- | --- |
| **Patient Age** | 51.3 (18.7) |
| **Age Category** |  |
| <50 | 148,179 (46%) |
| 50-75 | 141,110 (43%) |
| 75> | 35,489 (11%) |
| **Gender** |  |
| 2 | 127,389 (39%) |
| Female | 197,389 (61%) |
| **Smoker** |  |
| Never Smoker | 310,423 (96%) |
| Smoker | 14,355 (4.4%) |
| **Type 2 Diabetes Mellitus** | 68,196 (21%) |
| **Coronary Heart Disease** | 10,442 (3.2%) |
| **Stroke** | 18,690 (5.8%) |
| **Platelet Count** | 232.3 (62.0) |
| **Leukocyte Count** | 7.3 (2.9) |
| **Hematocrit** | 38.7 (9.2) |
| <sup>1</sup> Mean (SD); n (%) |  |

**Table 2.** Clinical characteristics for the UPHS study population divided into groups based on platelet count. Group 1 represents 100-200×10^9^/L, Group 2 (reference) represents 201-250×10^9^/L, Group 3 represents 251-300×10^9^/L, and Group 4 represents 301-450×10^9^/L.

| **Characteristic** | **1**<br>N = 105,860<sup>1</sup> | **2**<br>N = 107,272<sup>1</sup> | **3**<br>N = 67,865<sup>1</sup> | **4**<br>N = 43,747<sup>1</sup> | **p-value**<sup>2</sup> |
| --- | --- | --- | --- | --- | --- |
| **Patient Age** | 54.1 (19.2) | 50.6 (18.5) | 49.2 (18.0) | 49.5 (18.2) | <0.001 |
| **Gender** |  |  |  |  | <0.001 |
| FEMALE | 51,838 (49%) | 65,187 (61%) | 47,571 (70%) | 32,768 (75%) |  |
| MALE | 54,022 (51%) | 42,085 (39%) | 20,294 (30%) | 10,979 (25%) |  |
| **Smoker** | 4,217 (4.0%) | 4,516 (4.2%) | 3,221 (4.7%) | 2,395 (5.5%) | <0.001 |
| **Number of Comorbidities** |  |  |  |  | <0.001 |
| 0 | 76,437 (72%) | 81,923 (76%) | 51,621 (76%) | 31,263 (71%) |  |
| 1 | 24,390 (23%) | 21,761 (20%) | 14,105 (21%) | 10,742 (25%) |  |
| 2 | 4,475 (4.2%) | 3,235 (3.0%) | 1,924 (2.8%) | 1,555 (3.6%) |  |
| 3 | 558 (0.5%) | 353 (0.3%) | 215 (0.3%) | 187 (0.4%) |  |

<sup>1</sup> Mean (SD); n (%)
<sup>2</sup> Kruskal-Wallis rank sum test; Pearson's Chi-squared test

We observed a U-shaped relationship between platelet count and stroke risk for both ischemic and hemorrhagic stroke (**Fig. 3A-D**). These trends were consistent in men and women (**Fig. 3B-F**). We noted a closer correlation between platelet count and stroke at higher platelet counts. We hypothesized that stroke incidence at lower platelet counts might reflect concurrent illness, as opposed to a direct association with platelet count per se. Consistent with this idea, overall mortality was statistically higher at platelet count < 1.89 or > 2.69, with a more pronounced increase at lower platelet counts (**Fig. 3G**). This trend was consistent in men and women **(Fig. 3H-I).** Together, the epidemiologic association between platelet count and stroke risk spurred us to more deeply examine genetically determined causal relationships and mechanisms.

**Figure 3.**
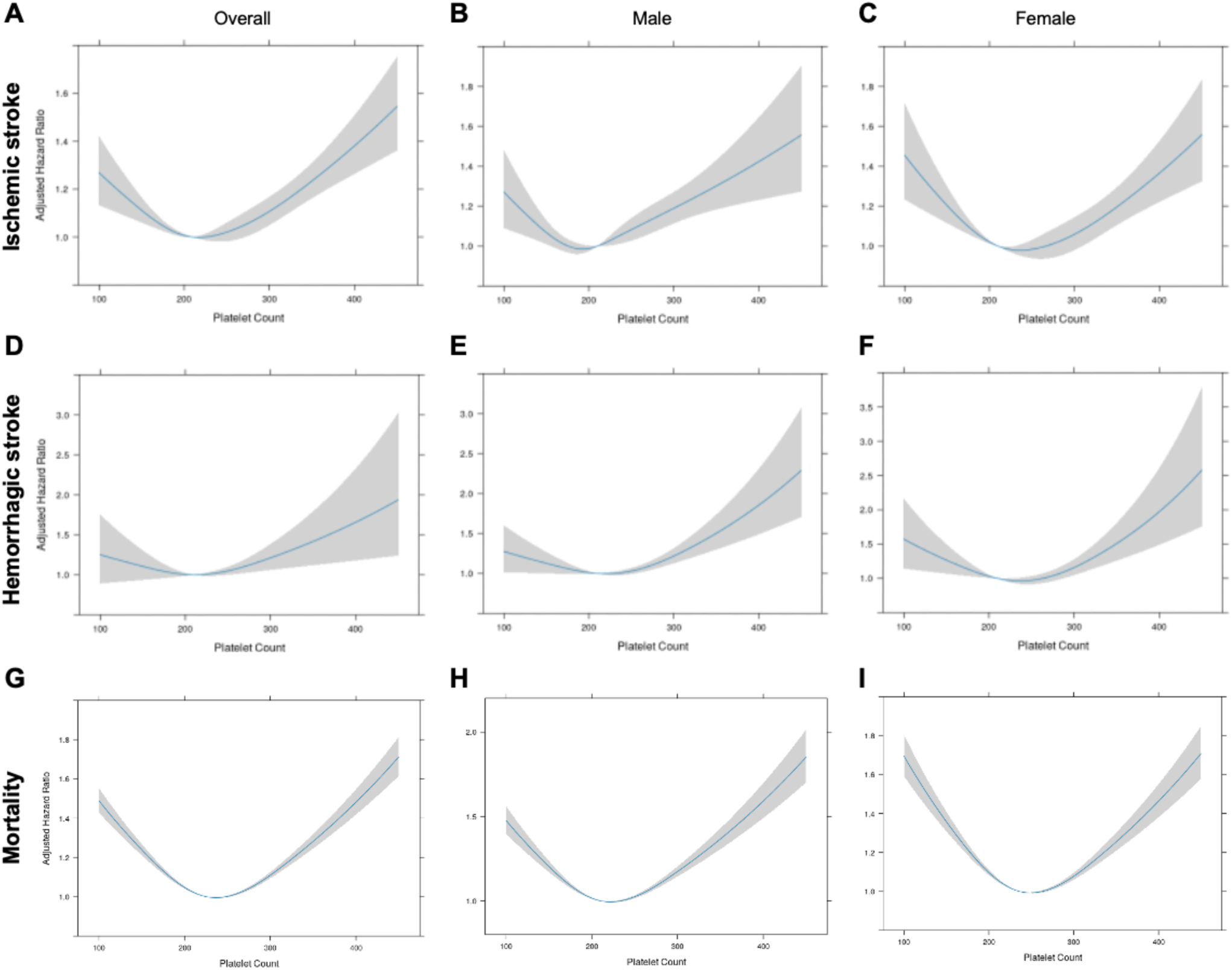
Platelet count variation is epidemiologically associated with stroke risk or mortality. A) Platelet count variation correlates with ischemic stroke risk. B) Platelet count variation correlates with ischemic stroke risk in males. C) Platelet count variation correlates with ischemic stroke risk in females. D) Platelet count variation correlates with hemorrhagic stroke risk. E) Platelet count variation correlates with hemorrhagic stroke risk in males. F) Platelet count variation correlates with hemorrhagic stroke risk in females. G) Platelet count variation correlates with mortality risk. H) Platelet count variation correlates with mortality risk in males. I) Platelet count variation correlates with mortality risk in females. Platelet count is represented in ×10^9^/L and shaded regions indicate 95% confidence intervals. Clinical characteristics for each group are listed in Table 2.

### Trait clustering reveals distinct genetic mechanisms and cell types linking platelet traits to stroke risk

Platelet counts can vary from heterogeneous biological mechanisms, including but not limited to genetically determined differences in bone marrow hematopoiesis, biased lineage-specific blood cell production, peripheral platelet activation with consumption, apoptotic cell clearance, generalized inflammation, and interactions with other cell types in circulation (e.g., endothelial cells)^26–29^. We reasoned that these varied mechanisms were integrated within platelet trait GWAS loci, and that this mechanistic admixture might account for the relatively weak effects of platelet count on stroke by MR (**Fig. 1**). By extension, we hypothesized that clustering platelet count GWAS loci based on trait associations would reveal subclusters with stronger direct impacts on stroke risk, leading to mechanistic, locus-specific, and translationally relevant insights.

The Noise-augmented von Mises-Fisher Mixture model (NAvMix) was designed to cluster variants according to cross-trait associations and has revealed novel mechanisms underlying complex disease biology^21^. To define platelet-related biological pathways and cell types linked to stroke risk, we applied NAvMix to cluster 1115 platelet-associated variants via locus-specific association with cardiometabolic, anthropometric, and autoimmune traits with established relationships to stroke risk and/or platelet biology (**Fig. 4A**). All variants were harmonized to effect alleles that positively influenced platelet count.

**Figure 4.**
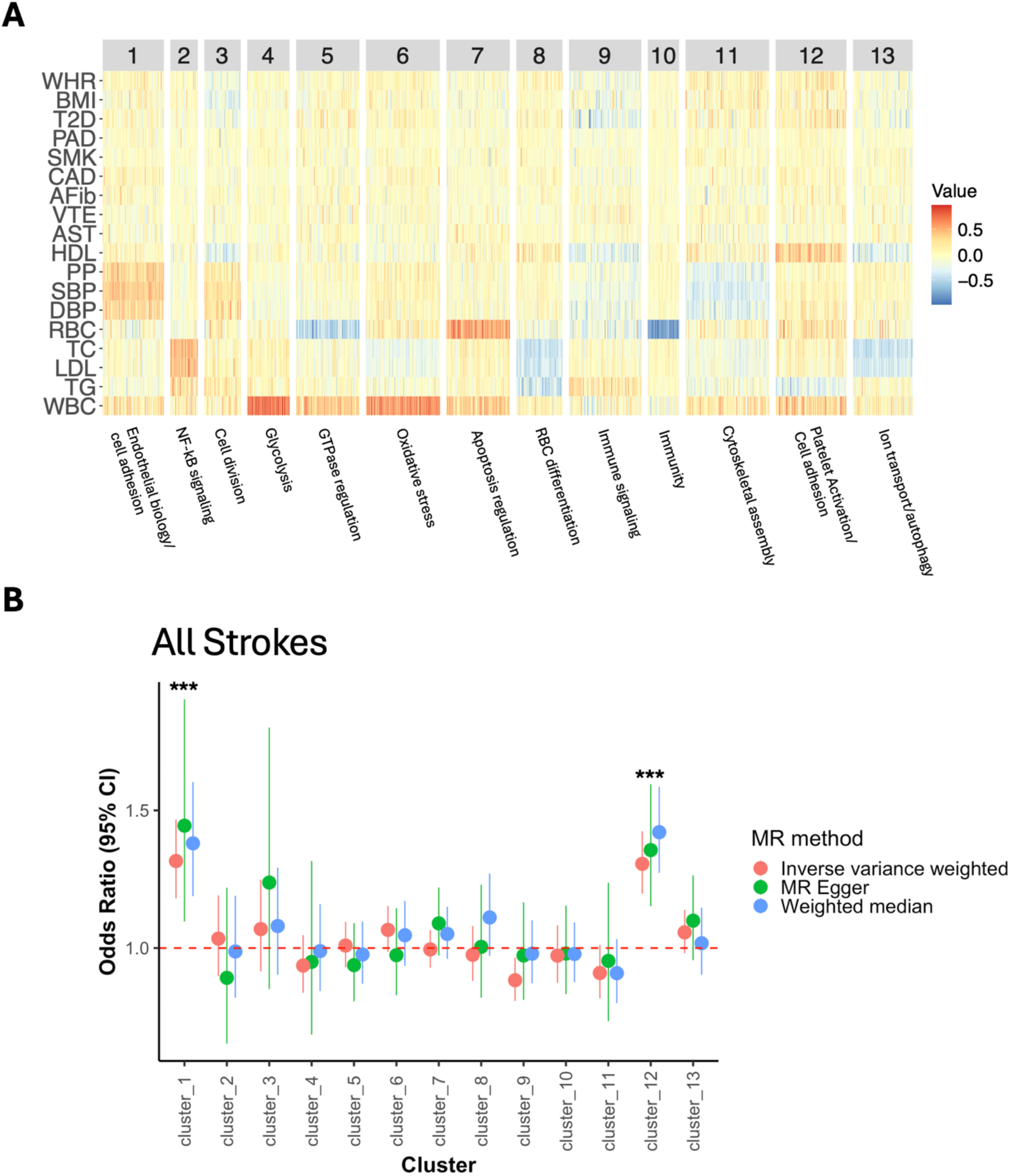
Clustering of PLT-associated genetic variants identifies subclusters of variants that impact stroke risk. (A) Heatmap of association estimates between platelet-associated variants and 18 stroke-related traits, grouped into 13 clusters by NAvMix. Colors reflect direction and magnitude of association with a trait within each cluster (red = positive, blue = negative). Clusters were annotated by manual curation of gene ontology (GO) terms. (B) Two-sample MR estimates (IVW, MR Egger, and weighted median) of the effect of each cluster on stroke risk. Clusters 1 and 12 show significant positive associations with stroke, which were consistent across MR methods. *p<0.05 by t-test after Bonferroni correction.

We identified 13 variant clusters based on trait association profiles (**Fig. 4A**). To ascertain functional and biological relevance for each cluster, we mapped genes to each cluster based on platelet expression quantitative trait loci (eQTLs)^30^. We then performed Gene Ontology (GO) analysis based on related genes and assigned cluster names based on prominent biological pathways^31^ (**Fig 4A**). These experiments confirmed that platelet GWAS loci reflect myriad cell types and biological processes, from hematopoietic cell development to inflammation to processes related to platelet activation.

We then used MR to identify SNP clusters that were causally related to stroke risk. Clusters 1 (“Endothelial cell biology/cell adhesion”) and 12 (“Platelet activation/cell adhesion”) significantly increased stroke risk, with consistent effect estimates across MR methods that exceeded the aggregated platelet count GWAS loci (OR∼1.31 [*P* < 10^-6^] in **Fig. 4B** vs OR 1.03 in **Fig. 1**). Cluster 1 variants were positively associated with blood pressure and WBC variation. GO enrichment analysis indicated links to endothelial cell biology (e.g., cell-matrix adhesion, epithelial cell-cell adhesion, wound healing, **Fig. 5A**). Cluster 12 variants were positively associated with HDL, RBC, WBC and cardiometabolic disease traits, and negatively associated with triglycerides. GO enrichment revealed biological associations GTPase enzyme activities, ribosomal function, and focal adhesion, (**Fig. 5B**). Pathway enrichment in cluster 12 was consistent with processes known to impact platelet activation and reactivity^32^. Of all platelet count clusters, cluster 12 was the most closely linked to platelet reactivity to epinephrine by MR, although we noted that platelet reactivity was a relatively underpowered outcome phenotype (OR 1.27 [0.92 to 1.75], *P* = 0.15)^13^ (**Supplemental Fig. 1**).

**Figure 5.**
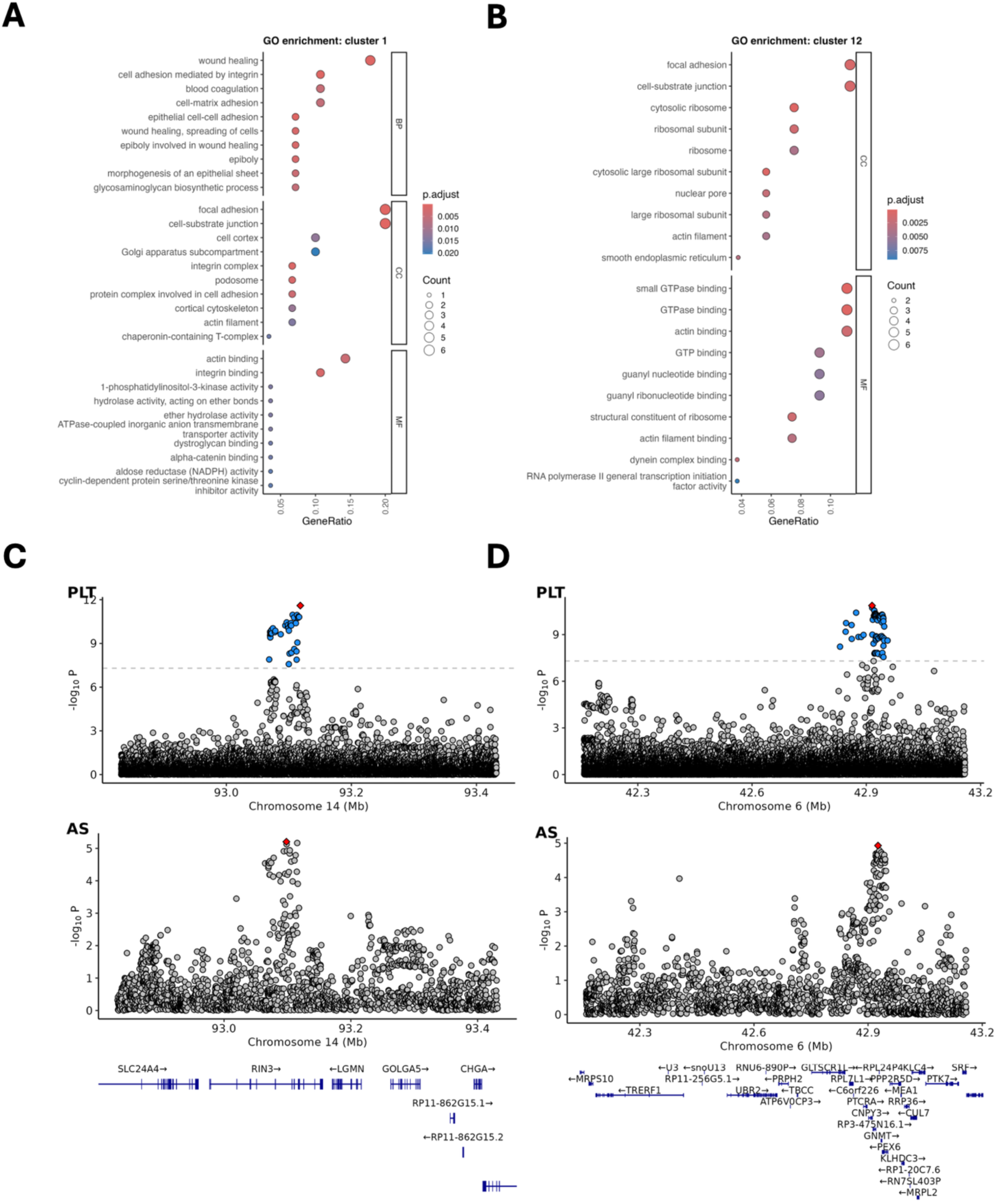
Platelet-specific loci and mechanisms increase stroke risk. (A) GO enrichment analysis of cluster 1 highlights pathways related to endothelial biology, including cell adhesion, wound healing, and epithelial morphogenesis. (B) GO enrichment analysis of cluster 12 highlights pathways related to platelet reactivity, including GTPase enzyme activity, ribosomal function, and cytoskeletal assembly. (C) LocusZoom plot of the RIN3 locus, encoding a Ras effector protein which is a binding partner to RAB5 small GTPases (D) LocusZoom plot of a representative locus from cluster 12 showing colocalized signals for platelet count and stroke at the *PEX6/PEX29* gene locus.

Having identified platelet trait-related SNP clusters highly predictive for stroke, we sought to identify loci linking platelet biology and stroke risk. We used genetic colocalization analyses to identify shared genetic architecture^33^. We defined a PP4>0.8 as statistical evidence suggesting genetic colocalization^17^. For example, SNPs near the *RIN3* gene locus showed evidence of colocalization (PP4>0.9, **Fig. 5C**). *RIN3* encodes a Ras effector protein that interacts with GTPases^34^. *RIN3* is highly expressed in platelets^35^ and linked to stroke risk^36^. In addition, this experiment indicated colocalized genetic signals near the *PEX6* and *PEX39* gene loci (**Fig. 5D**). These genes encode proteins essential for peroxisome biogenesis, protein import, and metabolic function, consistent with pathways that impact platelet reactivity^37^. These exemplary findings support our cluster annotations and illustrate how leveraging variant clustering can help define genetic loci that contribute to stroke risk through platelet-related mechanisms.

## Discussion

Genetic mechanisms underlying quantitative blood trait variation can relate to blood cell-intrinsic processes, including hematopoietic development and/or mature blood cell function, as well as systemic pathology that extrinsically regulated blood cell biology. A mix of cell types and processes can therefore modulate blood trait-related complex disease risks. Causal mechanisms linking blood traits to disease outcomes remain incompletely understood. We designed this study to interrogate epidemiologic and genetic links between blood cell traits and stroke risk. Our findings identify causal cell types and pathways linking blood trait variation to stroke, most notably implicating platelet trait loci in stroke pathophysiology through processes active in endothelial cells and platelets (**Fig. 4**). Our approach and highlighted loci provide novel insights into complex disease associations and could lead to novel drug targets.

Epidemiologic correlation supported a U-shaped relationship between platelet counts and stroke risk, as well as overall mortality (**Fig. 3**). This pattern was generally consistent with previously reported relationships in a smaller cohort^9^. Our cohort of subjects included hospitalized patients with potentially severe concurrent illness, which can decrease platelet counts through multiple comorbidities that reduce platelet production or increase consumption, including but not limited to sepsis, bone marrow suppression, drug-induced thrombocytopenia, hepatic disease, or subclinical thrombosis that were otherwise not reported or available in our clinical database^38^. These unrecognized comorbidities could explain increased mortality at lower platelet counts and a higher risk of mortality compared to stroke risk at lower platelet counts (**Fig. 3**). Conversely, elevated platelet counts may increase thrombosis and stroke risk via hypercoagulability, inflammation, or other pathophysiology.

Our two-sample MR analyses revealed that multiple blood cell types (platelets, erythrocytes, leukocytes) are genetically positively associated with increased stroke risk (albeit weakly, **Fig. 1**). MR-BMA was designed to disentangle highly correlated traits, and this was an ideal scenario to test the hypothesis that one or more blood lineages was the key driver for blood trait-stroke association. MR-BMA prioritized platelet related traits as causal factors for stroke, although HGB also ranked highly (**Fig. 2**). While we prioritized platelet trait variation and platelet biology in this study, our findings suggest that elevated HGB may also be a bona fide stroke risk factor, as has been previously suggested for other cardiovascular disease^39^.

Our clustering and colocalization analyses revealed shared genetic architecture between platelet count and stroke (**Fig. 5**). These results highlight the multitude of cell types and mechanisms that contribute to platelet trait variation, including integral processes to maintain endothelial cells and platelet homeostasis. Endothelial dysfunction is a major underlying feature of cardiovascular diseases, including stroke^40^. Platelet reactivity is influenced by endothelial dysfunction but involves disparate mechanisms and signaling pathways^41,42^. *RIN3* is associated with GTPase activity and platelet reactivity, and peroxisome biogenesis represents a novel pathway linking platelet biology and stroke risk. We anticipate that future interrogation of these and other implicated loci will reveal novel therapeutic targets in stroke prevention and therapy.

During this study, we noted that relatively underpowered stroke GWAS datasets may have precluded more robust results. All GWAS used in this study were biased toward individuals of European ancestry, which may limit generalizability to other ancestries. These findings underscore the need for more robust and well-annotated multi-ancestry stroke GWAS in the future. While NAvMix trait selection was hypothesis-driven, future studies may also incorporate other relevant traits relating platelet biology to stroke risk.

Our findings provide definitive epidemiological and genetic evidence that increased platelet count heightens stroke risk and begins to elucidate related biological mechanisms. Integrating MR, MVMR, and MR-BMA with variant clustering and downstream pathway analyses helped determine cell types and platelet-related pathways that contribute to stroke risk. Our findings will inform the development of more targeted stroke prevention and treatment strategies.

## Methods

### Platelet and cardiovascular risk epidemiology

We identified 324,677 participants with platelet count measurements within the University of Pennsylvania Health System (UPHS), including inpatients and outpatients. All studies were approved by the University of Pennsylvania Institute Review Board (IRB). We excluded 4,759 (1.4%) with thrombocytopenia of whom 2,828 (59%) were male. We excluded 4,973 (1.5%) with thrombocytosis of whom 1,888 (38%) were male. The mean follow-up time was 9.0 ± 4 years among participants, during which 38,174 (12%) died. The mean change in platelet count was 2.1 ± 90 among participants who had another platelet count obtained within the study period, which occurred not more than 182 days after the initial platelet measurement (n = 38). Adjusted hazard ratios for stroke or overall mortality were based on platelet counts from the UPHS database adjusted for age, gender, leukocyte count, hematocrit, and comorbidities using Cox proportional hazard regression.

### GWAS collection

We used publicly available GWAS summary statistics for the exposure traits related to 15 quantitative blood measurements^23^ and stroke^22^. Summary statistics were originally or lifted^43^ to represent genome build hg19. Traits related to stroke included all strokes (AS, N = 110,182), any ischemic stroke (AIS, N = 86,668), cardioembolic stroke (CES, N = 12,790), large artery stroke (LAS, N = 9,219), and small vessel stroke (SVS, N = 13,620)^22^. Stroke type was defined by established medical criteria and/or neuroimaging results from medical records when available, as described in the original GWAS. Quantitative blood traits included genetic associations of red blood cell, white blood cell, and platelet phenotypes (N = 563,085)^23^. For clustering analyses, we additionally included GWAS summary statistics for 18 cardiometabolic, anthropometric, autoimmune, and lipid related trait, including smoking (SMK, N = 2,669,029)^44^, blood pressure [systolic (SBP), diastolic (DBP), and pulse pressure (PP) (N = 750,000 for all blood pressure traits)^45^, lipids [high density lipoprotein (HDL), low density lipoprotein (LDL), triglyceride level (TG), and total cholesterol (TC) (N = 1,320,000 for all lipid measurement traits)^46^, type 2 diabetes (T2D, N = 1,528,967)^47^, atrial fibrillation (AFib, N = 1,244,730)^48^, coronary artery disease (CAD, N = 296525)^49^, peripheral artery disease (PAD, N = 174,992)^50^, venous thromboembolism (VTE, N = 1,500,861)^51^, asthma (AST, N = 1,800,785)^52^, body mass index (BMI) and waist-hip-ratio (WHR) (N = 694,649 for both)^53^.

### Genetic variant selection and instrumental variable creation

We selected genetic instrumental variables by filtering GWAS summary statistics for linkage-independent SNPs common to exposure, outcome, and/or related risk factors. We focused on European ancestry data, although some datasets included cross-ancestry data in the clustering analyses to optimize power. We used R v4.2.3^54^, TwoSampleMR^55^, and the ieugwasr package^56^ to clump SNPs meeting genome-wide significance for each exposure phenotype, retaining independent SNPs (EUR r^2^ < 0.01) as instruments.

### Mendelian randomization analyses

We used TwoSampleMR to conduct MR analyses^55^. For MR-BMA analyses, we used the summary_mvMR_SSS and summary_mvMR_BF packages^20^. We estimated causal effects using the inverse-variance weighted (IVW), weighted median, and MR-Egger methods. IVW assumes all instruments are valid and provides the greatest statistical power. The weighted median method provides a valid estimate if at least 50% of the instruments are valid, while MR-Egger accounts for potential directional pleiotropy through its intercept test^57^. Effect estimates are reported per standard deviation (SD) increase in the exposure trait. Since exposures generally represented continuous variables (e.g., quantitative blood counts), outcome estimates reflect the effect of a 1 SD unit increase in exposure. For context, this is ∼1 g/dL HGB^58^. In all experiments, statistical significance was defined as *P* < 0.05 and 95% confidence intervals were calculated as 1.96 × standard error (SE). For MR-BMA analyses, we allowed up to five exposures per model and ran 100,000 stochastic search iterations to explore the model space.

### SNP cluster definition

We used the navmix package^21^ to perform clustering on platelet count associated SNPs. We selected 18 related cardiometabolic, anthropometric, autoimmune, lipid, and blood traits, based on known or suspected relevance to platelet biology, to capture groups reflecting biological processes underlying stroke risk. We allowed up to 20 clusters in our NAvMix experiments based on the lowest Bayesian Information Criterion (BIC) score. In these analyses, we considered 1115 linkage-independent variants associated with platelet count (*P* < 5×10⁻⁸, clumped at *r²* < 0.01).

### Pathway analyses

To identify eQTLs, we first identified all SNPs in high linkage disequilibrium (r^2^ < 0.8) with the 1115 platelet count-associated SNPs used in NavMix clustering analyses. These SNPs were mapped to genes using platelet eQTL data^30^. Gene ontology (GO) enrichment analysis was performed using the clusterProfiler package^59^ and visualized using the enrichplot package^60^. Clusters were annotated through manual curation and summarization of GO enrichment terms.

### Colocalization analyses

We performed colocalization analyses using the coloc package^33^, assuming a single causal variant within a 250 kb window. This analysis included platelet count–associated instruments from the clustering analysis as well as all SNPs in high linkage disequilibrium (r^2^ < 0.8) with these instruments. Loci with strong evidence of colocalization (posterior probability for a shared causal variant, PP4 > 0.8) were visualized with locuszoom^61^.

## Data Availability

All statistical analyses and output produced in the present study are available upon reasonable request to the authors.

## Ethics and Data Use Approval

The IRB of the University of Pennsylvania gave ethical approval for epidemiologic studies. This study used publicly available human GWAS summary statistics. Data use committee approval was required to access some data (dbGap Project #31950).

## Acknowledgements

This study was supported by the National Institutes of Health (NHLBI K99 HL156052 to CST).

## Conflicts of Interest

S.M.D. receives research support from RenalytixAIm, in-kind research support from Novo Nordisk and Amgen, and consulting fees from Tourmaline Bio, all outside the scope of the current project. Other authors declare no conflict of interests.

