## Supplementary material for "Genetically determined platelet traits impact stroke risk through multiple mechanisms and cell types": Supp Info

Kim Ha[1,2], Katherine Hartmann[3], Renae L Judy[4], Michael Levin[5,6], Scott Damrauer[4,5], Christopher S Thom[1,7,*]
[1] Division of Neonatology, Children's Hospital of Philadelphia, Philadelphia, PA, USA
[2] Cell and Molecular Biology Graduate Group, University of Pennsylvania Perelman School of Medicine, Philadelphia, PA, USA
[3] Department of Radiology, University of Pennsylvania Perelman School of Medicine, Philadelphia, PA, USA
[4] Department of Surgery, University of Pennsylvania Perelman School of Medicine, Philadelphia, PA, USA
[5] Corporal Michael J. Crescenz VA Medical Center, Philadelphia, PA, USA
[6] Division of Cardiovascular Medicine, University of Pennsylvania Perelman School of Medicine, Philadelphia, PA, USA
[7] Department of Pediatrics, University of Pennsylvania Perelman School of Medicine, Philadelphia, PA, USA

[*]Correspondence:
Christopher S Thom
10-052 Colket Translational Research Building
3501 Civic Center Blvd.
Philadelphia, PA 19104


Key Words: Platelet, Mendelian randomization, GWAS, Genetics

**Supplementary Information**

**Supplemental Figures**

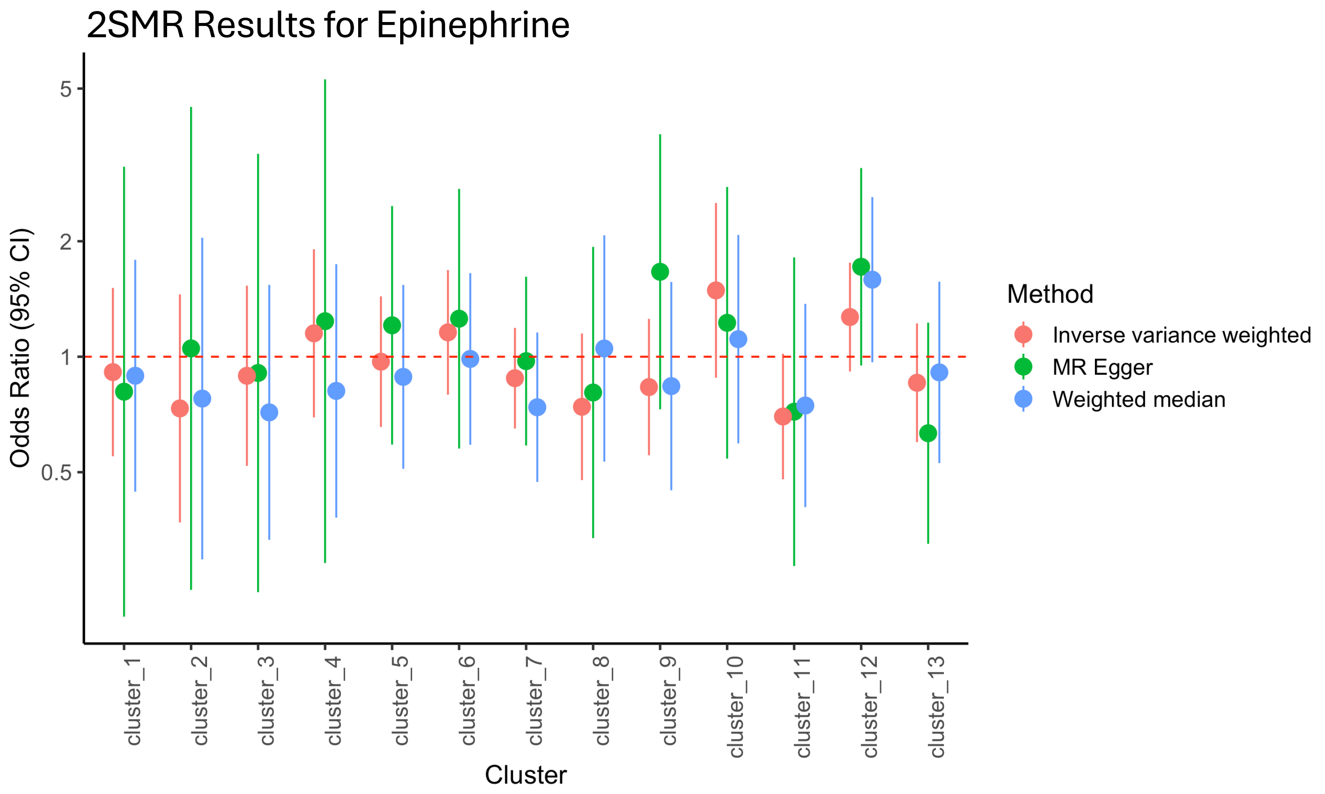


**Supplemental Figure 1**. Two sample MR effect estimates for 13 PLT GWAS-associated clusters on epinephrine-induced platelet aggregation.
